# Strategies to establish a biologically defined Parkinson’s disease cohort prior to the onset of clinically significant motor symptoms

**DOI:** 10.64898/2026.10.26.26366535

**Authors:** Ethan G. Brown, Helena Blumenau, David-Erick Lafontant, Anuprita R. Nair, Noah Johnson, Roy N. Alcalay, Lana M. Chahine, Christopher S. Coffey, Roseanne D. Dobkin, Tatiana Foroud, Douglas Galasko, Kalpana Merchant, Brit Mollenhauer, Kathleen L. Poston, Andrew Siderowf, Caroline M. Tanner, Daniel Weintraub, Kenneth Marek, Tanya Simuni, Thomas F. Tropea, the Parkinson’s Precision Medicine Initiative

**Author notes:** Corresponding Author: Ethan G. Brown, MD, University of California, San Francisco, 1651 4th Street | Level 3 SW Academic Offices, 364A, Box #1838 Weill Institute Building | San Francisco, CA 94158. These authors contributed equally (Tanya Simuni, Thomas F. Tropea).

## Abstract

Identifying individuals at risk for neurodegeneration is a priority for clinical trials. Parkinson’s Precision Medicine Initiative (PPMI) deploys strategies to enroll individuals before typical clinical Parkinson’s disease symptoms: recruiting polysomnogram-confirmed REM sleep behavior disorder (RBD) and central identification of hyposmia via smell testing. Groups were divided into RBD with and without hyposmia (RBD+Hyposmia, RBD-Hyposmia) and hyposmia with and without self-reported dream enactment behavior (Hyposmia+DEB, Hyposmia-DEB). Participants underwent clinical and biomarker testing including CSF alpha-synuclein seed amplification assay (CSFaSynSAA) and dopamine transporter imaging. While all enrollment strategies showed robust enrichment for CSFaSynSAA positivity, RBD+Hyposmia and Hyposmia+DEB were more likely CSFaSynSAA+ (94% and 85%) vs RBD-Hyposmia (40%) and Hyposmia-DEB (55%). Among CSFaSynSAA+, RBD+Hyposmia and Hyposmia+DEB had more mood, autonomic, and cognitive symptoms than Hyposmia-DEB. PPMI identified a large biologically-defined early neuronal synucleinopathy cohort and demonstrated biomarker and multi-domain clinical heterogeneity within CSFaSynSAA+ individuals prior to symptom onset.

## Introduction

Neurodegeneration begins years prior to a clinical diagnosis of Parkinson’s disease (PD).^1,2^ Recent availability of the cerebrospinal fluid alpha-synuclein seed amplification assay (CSFasynSAA) coupled with dopamine transporter (DAT) imaging to detect early reduction in DAT binding have enabled the identification of individuals with neuronal synuclein disease (NSD) prior to typical onset of PD motor symptoms.^3^ Identifying people early in the neurodegenerative process is important for therapeutic studies attempting to prevent disease progression and for observational studies seeking to understand the pathophysiology of early disease. However, current biomarker tests are not amenable to large-scale screening, so additional screening or enrichment methods are required.

The Parkinson’s Precision Medicine Initiative (PPMI) study has enrolled > 2000 prodromal individuals without a clinical diagnosis of PD based on their biomarker characterization. Participants are eligible for biomarker assessment based on either a diagnosis of rapid eye movement sleep behavior disorder (RBD) or hyposmia according to smell identification testing.^4^ Isolated RBD, a condition characterized by dream enactment behavior (DEB), is the most specific prodromal condition, but only occurs prior to diagnosis in ∼25% of people with PD.^5,6^ Furthermore, definitive diagnosis of RBD requires detection of REM sleep without atonia on polysomnography (PSG), which can only be done at specialized centers. Hyposmia is present in approximately 75-90% of individuals with PD and can occur years prior to any other symptoms.^7,8^ Other prodromal symptoms, such as constipation and mood disorders, are less specific. Identifying people with combinations of these symptoms in whom additional biological characterization through biochemical or imaging biomarkers is warranted can be a useful enrichment strategy to improve research study conduct.^9–11^

Our prior work demonstrated that targeted biomarker characterization could be implemented to enrich a prodromal cohort for DAT deficit and positive CSFaSynSAA through staged screening (i.e., smell testing to identify hyposmia followed by DAT imaging).^3,4^ The objective of this paper is to (a) investigate NSD prevalence among people with iRBD and/or hyposmia enrolled in PPMI and (b) characterize biological and clinical subgroups of people enrolled with prodromal symptoms.

## Methods

### Population and group definition

Data were from the PPMI, a multicenter longitudinal cohort study. Participants enrolled after 2020 in the Prodromal Cohort of PPMI were included in this analysis.^12^ Full details of the PPMI protocol can be found elsewhere.^13,14^ Briefly, participants were eligible if they were at least 60 years old, did not have a clinical diagnosis of PD, dementia with Lewy bodies (DLB), multiple system atrophy (MSA), or another neurodegenerative disease (Figure 1). Additionally, participants either had (a) RBD with polysomnographic confirmation reported by the site investigator (RBD-PSG) or (b) hyposmia determined by the University of Pennsylvania Smell Identification Test (UPSIT) and DAT binding that was less than or equal to 100% of expected for age and sex.^13,15^ Hyposmia was defined as ≤15^th^ percentile, using normative data derived from PPMI.^16^ DAT imaging was conducted using [123I]FP-CIT SPECT.^12,13^ The lowest putamen specific binding ratio was determined and compared to age- and sex-adjusted expected values for eligibility. Participants were excluded from this analysis if they were enrolled prior to 2020 or had parkinsonism without a clinical diagnosis of PD or dementia in the opinion of the investigator on the baseline exam. All participants signed informed consent and the study has been approved by the WCG Institutional Review Board.

**Figure 1:**
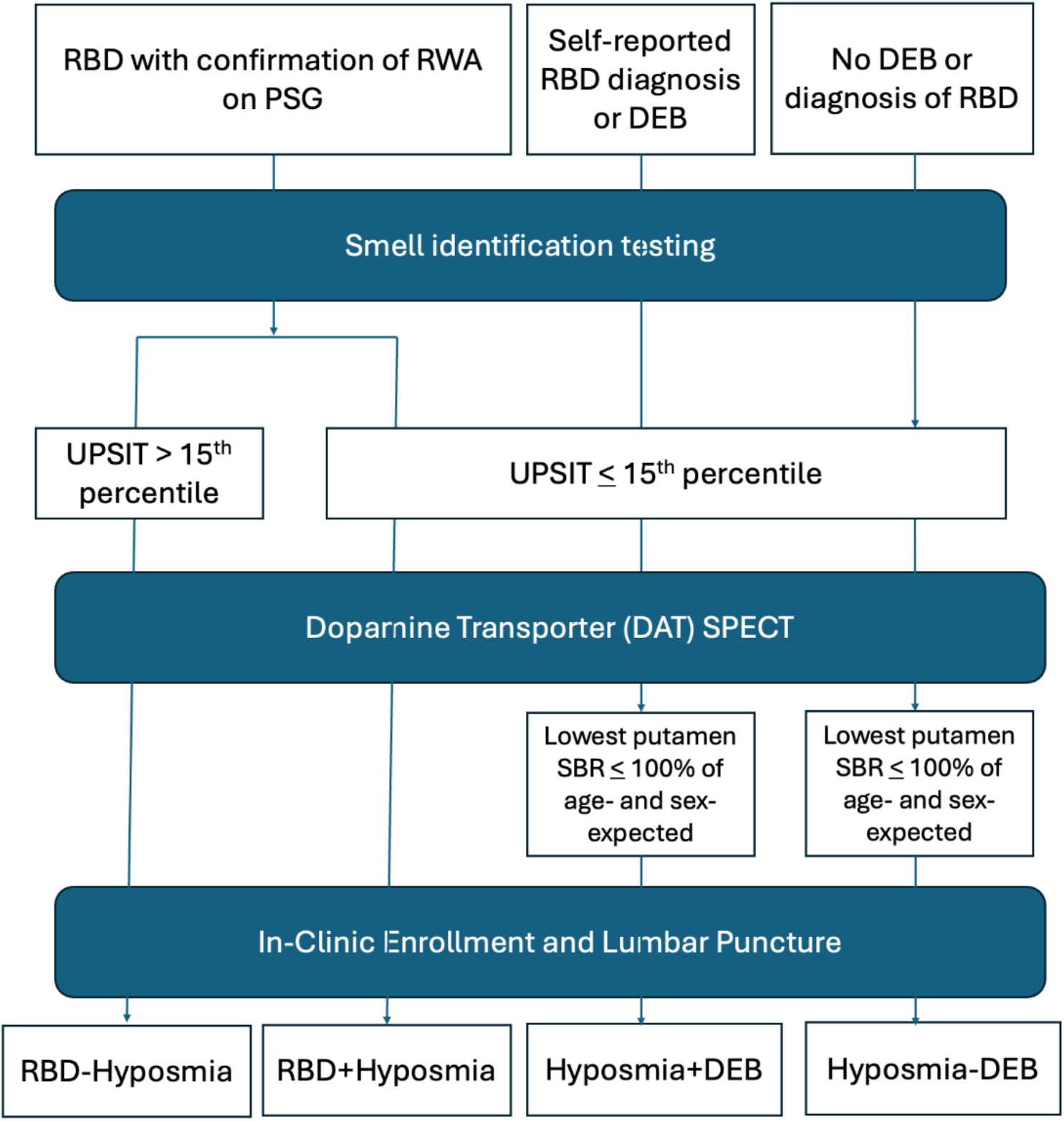
Flow diagram of participant eligibility and subgroup definition. RBD: REM sleep behavior disorder. RWA: REM sleep without atonia. PSG: polysomnography. DEB: dream enactment behavior. SBR: Specific binding ratio.

### Assessments

All assessments were conducted as part of PPMI in-person study activities. People identified as RBD via PSG testing were also administered an UPSIT and underwent DAT imaging, although the results of these tests did not determine eligibility for this group. People identified as hyposmic were given questionnaires at the time of UPSIT completion to ascertain a prior RBD diagnosis or whether they experienced DEB. DEB was assessed with the following question, adapted from a single-item screen:^17^ “Some people act out dreams while they sleep. They may punch, kick, yell or even fall out of bed. Have you been told you act out your dreams? Or do you suspect you many do this?” Questionnaire responses were not part of eligibility determination. Longitudinal assessments utilized standardized rating scales to characterize motor and non-motor symptoms. For this analysis, we included assessments of motor, cognitive, mood, and autonomic domains from baseline or screening visits as follows: Movement Disorders Society-Unified Parkinson’s Disease Rating Scale (MDS-UPDRS) parts I, II, and III as global measures of motor and non-motor symptoms,^18^ Montreal Cognitive Assessment (MoCA),^19^ cognition characterization based on the opinion of the investigator (i.e., normal cognition, mild cognitive impairment [MCI], or dementia), and a cognitive summary score derived based on neuropsychological tests as measures of cognition.^20^ The 15-item Geriatric Depression Scale (GDS)^21^ and the State-Trait Anxiety Inventory^22^ were measures of mood and anxiety. The Scales for Outcomes in Parkinson’s Disease – Autonomic Dysfunction (SCOPA-AUT)^23^ measures autonomic symptoms. Longitudinal DAT imaging and MR imaging was also acquired in all participants. Additional assessments conducted include the neurologic exam, blood tests, and lumbar puncture.^13^ CSFasynSAA was conducted on all cases at study entry.^24^

### Subgroup Definition

To explore whether the presence of RBD, DEB, or hyposmia alone, or in combination, at enrollment is associated with biomarker and clinical characteristics in the prodromal stage, we analyzed the data according to the following subgroups at baseline (Figure 1):

- **RBD+Hyposmia**: participants eligible based on RBD-PSG with hyposmia (< 15^th^ percentile) on UPSIT.
- **RBD-Hyposmia**: participants eligible based on RBD-PSG who did not have hyposmia (> 15^th^ percentile) on UPSIT at the baseline visit.
- **Hyposmia+DEB**: participants who were eligible based on hyposmia and DAT binding <100% who reported DEB or a diagnosis of RBD on the screening questionnaire (most often without PSG confirmation).
- **Hyposmia-DEB**: participants who were eligible based on hyposmia and DAT binding who did not endorse either DEB or diagnosis of RBD on the screening questionnaire.

Across all groups, we defined NSD as participants with a positive CSFaSynSAA at baseline.^25^ People with a negative or inconclusive CSFaSynSAA were considered not NSD.

In general, the positive CSFaSynSAA participants were enrolled at stage 2 of the NSD-ISS. They demonstrate aSyn pathology based on any validated biomarker with symptoms – such as hyposmia or DEB – but without functional impairment or evidence of typical PD.^26^ NSD stage 2 is in turn subdivided into Stage 2A if the person does not demonstrate a dopaminergic deficit on any validated biomarker (here DAT) and stage 2B if they do demonstrate a dopaminergic deficit.

### Statistical analyses

Demographic information and clinical characteristics were summarized across subgroups using descriptive statistics. Comparisons across groups were performed using global model-based tests of subgroup effects. Logistic regression models were used for categorical outcomes and general linear models for continuous outcomes. All models were adjusted for age and sex except scores that already accounted for these variables (i.e., UPSIT percentile, age- and sex-adjusted DAT (percent expected), and the cognitive summary score). Residual normality was assessed for all continuous outcomes. When model assumptions were not met, appropriate transformations (square root, square, or logarithmic) were applied. If assumptions remained violated, rank-based analyses were used. Statistical significance was evaluated using Benjamini-Hochberg (B-H) false discovery rate (FDR) procedure. For global tests, comparisons with p-values below the BH threshold (α = 0.033) were considered statistically significant. For outcomes with statistically significant global subgroup effects after B-H adjustment, pairwise subgroup comparisons were conducted using the same modeling approach and outcome transformations as in the corresponding global models. A separate B-H procedure was applied to the set of pairwise p-values, with significance defined as p-values below the corresponding BH threshold (α = 0.040).

## Data availability

Data used in the preparation of this article were obtained on June 9, 2025 from the PPMI database (https://www.ppmi-info.org/access-data-specimens/download-data), RRID:SCR_006431. For up-to-date information on the study, visit http://www.ppmi-info.org. This analysis used DAT and CSFaSynSAA results for prodromal participants, obtained from PPMI upon request after approval by the PPMI Data Access Committee.

## Results

### Demographic and Clinical Summary

Participants were enrolled between October 2020 and June 2025. Participants eligible due to the presence of RBD were recruited from 55 international study sites. Participants recruited based on hyposmia were recruited through smell testing implemented remotely in the United States. Ultimately, 1,652 were included in this analysis. After enrollment, participants were assigned to an analytic subgroup: RBD+Hyposmia N=239 (14% of total enrollment), RBD-Hyposmia N=147 (9%), Hyposmia+DEB N=518 (31%), and Hyposmia-DEB N=748 (45%).

Demographic characteristics across groups are shown in Table 1. The number of participants enrolled with a PSG-confirmed RBD diagnosis (N=386, the RBD+Hyposmia and RBD-Hyposmia groups) were lower than individuals enrolled based on hyposmia (n = 1,266, the Hyposmia+DEB and Hyposmia-DEB groups). A higher proportion of people identified based on hyposmia were female compared to the RBD groups (58% in the Hyposmia+DEB/Hyposmia-DEB groups vs 22% in the RBD+Hyposmia/RBD-Hyposmia groups). Individuals enrolled with Hyposmia+DEB were slightly older than other groups (mean age of Hyposmia+DEB 69.2 vs RBD+Hyposmia 68.0, RBD-Hyposmia 67.3, and Hyposmia-DEB 67.6).

**Table 1:**
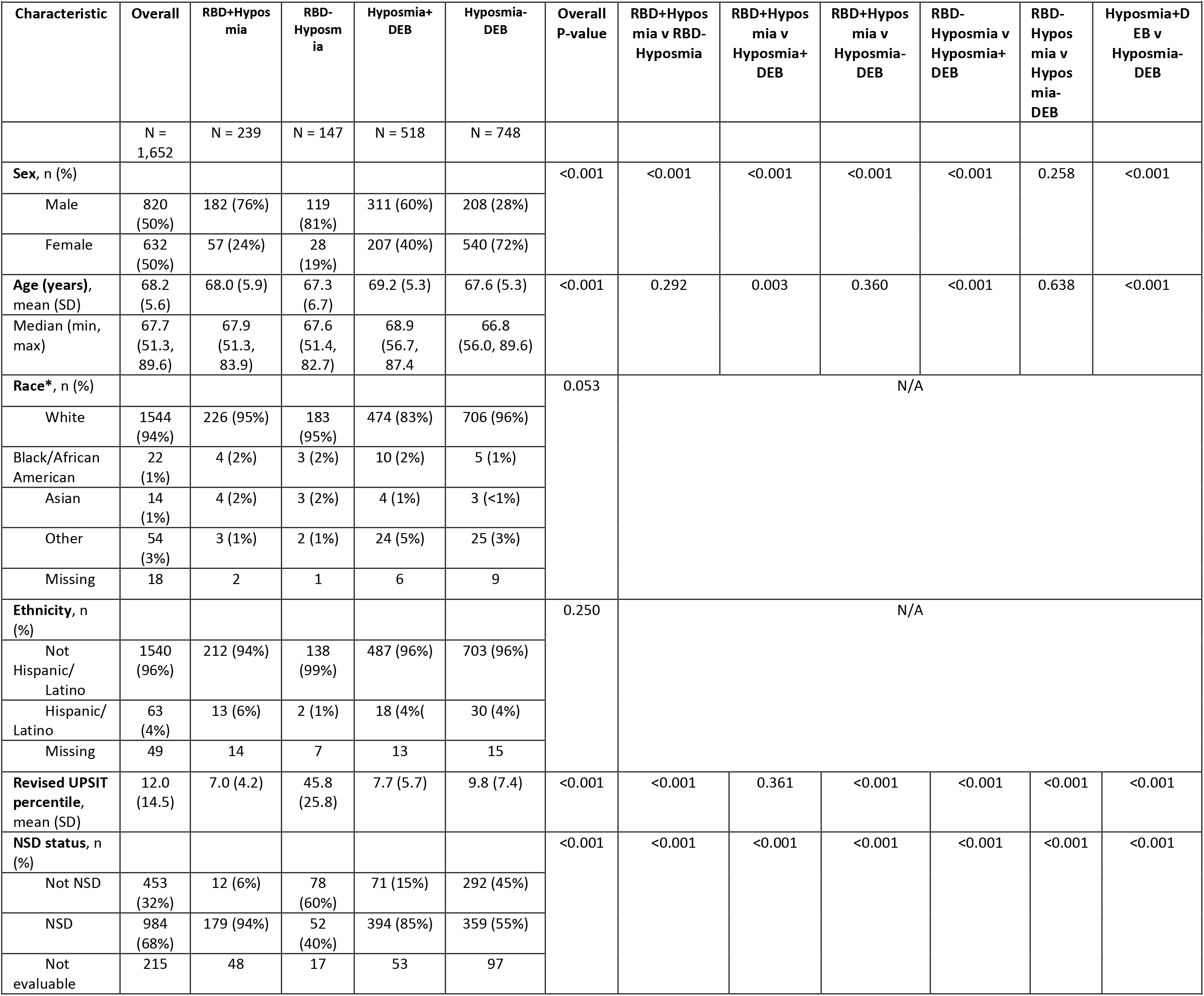
Clinical characteristics and NSD positivity in people with different clinical phenotypes of prodromal PD.

### NSD prevalence varies with prodromal phenotype

The frequency of NSD+ (i.e., positive CSFaSynSAA) varied by prodromal subgroup (Figure 2). Among people with RBD+Hyposmia, 93.7% were NSD+, while among the RBD-Hyposmia group only 40%. Were NSD+. Among people with Hyposmia+DEB, 84.7% were NSD+. A lower proportion of people with Hyposmia-DEB were NSD+ (55.1%).

**Figure 2:**
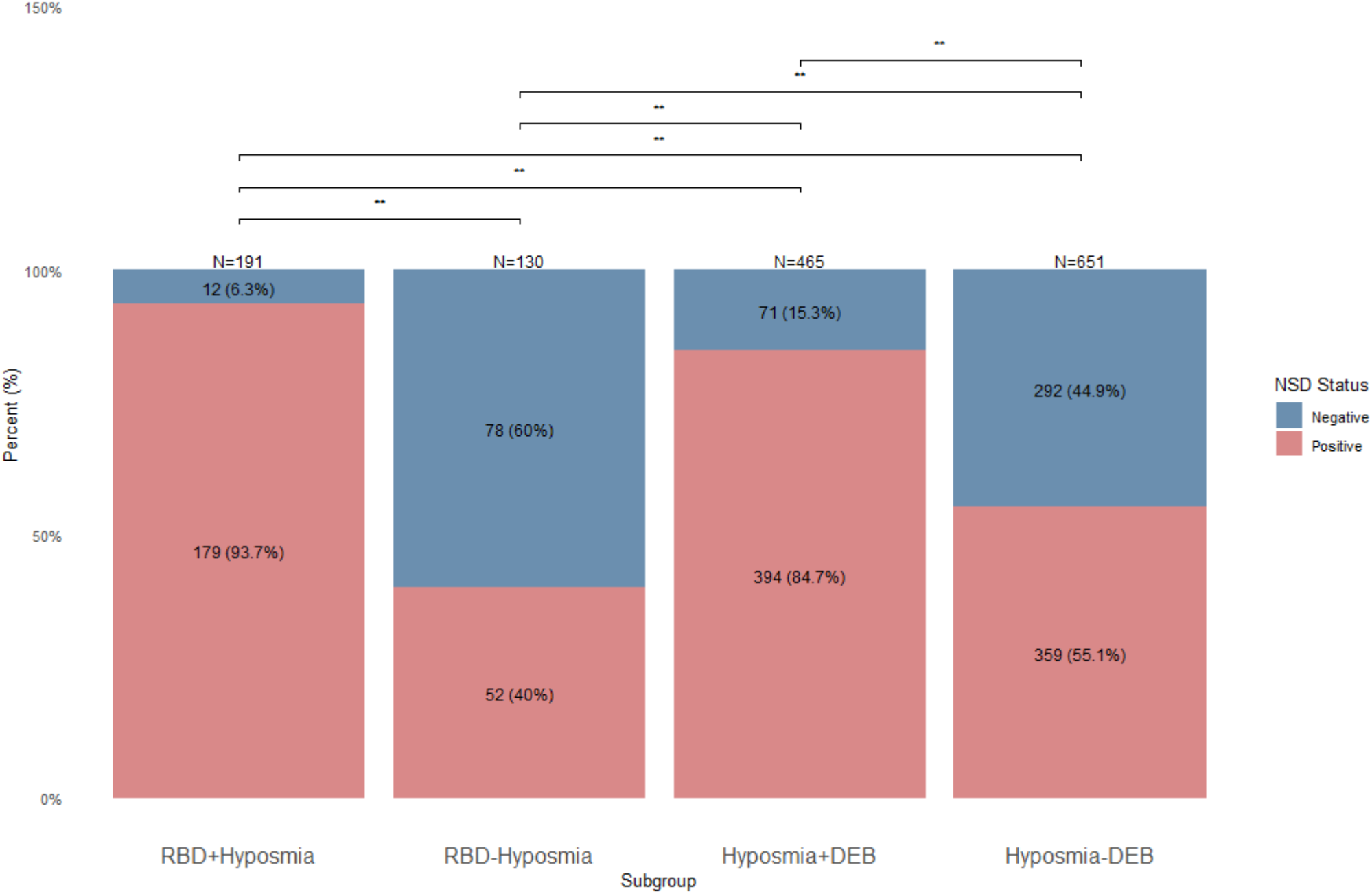
NSD is most prevalent among people with RBD-PSG or dream enactment behavior with hyposmia . Stacked bar-graph representing different proportions of NSD positivity (i.e., cerebrospinal fluid alpha-synuclein seed amplification assay) across prodromal subgroups. Participants are identified either based on REM sleep behavior disorder confirmed on polysomnography (RBD-PSG) or through scoring less than the 15^th^ percentile on the University of Pennsylvania Smell Identification Test (UPSIT). RBD+Hyposmia: Participants identified based on RBD-PSG with hyposmia (UPSIT ≤ 15^th^ percentile). RBD-Hyposmia: Participants identified based on RBD-PSG without hyposmia. Hyposmia+DEB: Participants identified based on hyposmia who endorse dream-enactment behavior. Hyposmia-DEB: Participants identified based on hyposmia who deny dream-enactment behavior.

### Clinical features between NSD and non-NSD participants

Demographic and clinical characteristics were compared between NSD+ versus NSD-within analytic subgroups (**Table 2**). Within each subgroup, lower UPSIT percentile was associated with NSD+ compared with NSD-. NSD+ versus NSD-groups were older among RBD-Hyposmia, Hyposmia+DEB, and Hyposmia-DEB and more often male among Hyposmia+DEB and Hyposmia-DEB. NSD+ versus NSD-groups had lower age and sex-corrected DAT percentile among RBD+Hyposmia, DEB+Hyposmia and Hyposmia-DEB. Among people with DEB+Hyposmia, NSD+ was associated with higher total score on the RBD Questionnaire (p<0.001) compared to NSD-individuals. On clinical assessments, among NSD+ participants compared with NSD-, we observed lower MDS-UPDRS Part I (RBD+Hyposmia, Hyposmia-DEB), MDS-UPDRS Part II (RBD-Hyposmia, Hyposmia+DEB), MDS-UPDRS Total (RBD+Hyposmia, Hyposmia+DEB), SCOPA-AUT (RBD+Hyposmia, Hyposmia-DEB), GDS (Hyposmia-DEB), STAI (RBD+Hyposmia), and higher MoCA (Hyposmia+DEB) scores.

**Table 2:** Differences between NSD- and NSD+ across groups.

|  | RBD+Hyposmia |  |  | RBD-Hyposmia |  |  | Hyposmia+DEB |  |  | Hyposmia-DEB |  |  |
| --- | --- | --- | --- | --- | --- | --- | --- | --- | --- | --- | --- | --- |
|  | NSD-<br>(N = 12) | NSD+<br>(N=179) | P-value | NSD-<br>(N = 78) | NSD+<br>(N=52) | P-value | NSD-<br>(N = 71) | NSD+<br>(N=394) | P-value | NSD-<br>(N = 292) | NSD+<br>(N=359) | P-value |
| **Sex, n (%)** |  |  | 0.538 |  |  | 0.220 |  |  | <0.001 |  |  | <0.001 |
| Male | 10 (83%) | 135 (75%) |  | 59 (76%) | 44 (85%) |  | 25 (32%) | 251 (64%) |  | 58 (20%) | 129 (36%) |  |
| Female | 2 (17%) | 44 (25%) |  | 19 (24%) | 8 (15%) |  | 46 (65%) | 143 (36%) |  | 234 (80%) | 230 (64%) |  |
| **Age (years), mean (SD)** | 65.4<br>(3.9) | 67.9<br>(6.1) | 0.164 | 65.8<br>(6.5) | 68.9<br>(6.5) | 0.008 | 66.2<br>(4.9) | 69.7<br>(5.2) | <0.001* | 66.2<br>(4.9) | 68.4<br>(5.3) | <0.001* |
| **Revised UPSIT percentile, mean (SD)** | 9.9<br>(4.1) | 6.9<br>(4.2) | 0.020* | 55.8<br>(24.2) | 32.0<br>(20.0) | <0.001* | 10.8<br>(7.6) | 7.2<br>(4.9) | <0.001* | 12.9<br>(8.6) | 7.4<br>(5.2) | <0.001* |
| **% Age/sex-expected low putamen, mean (SD)** | 1.05<br>(0.21) | 0.77<br>(0.27) | <0.001 | 0.88<br>(0.23) | 0.82<br>(0.24) | 0.136 | 0.73<br>(0.22) | 0.67 (0.20) | 0.003* | 0.78<br>(0.14) | 0.70<br>(0.18) | <0.001* |
| Missing | 0 | 6 |  | 4 | 1 |  | 0 | 26 |  | 0 | 21 |  |
| **MDS-UPDRS part I, mean (SD)** | 11.9<br>(7.0) | 7.7<br>(5.8) | 0.017* | 8.5<br>(6.4) | 8.0<br>(4.7) | 0.704** | 8.6<br>(5.7) | 7.7<br>(5.0) | 0.711* | 6.3<br>(4.9) | 5.3<br>(4.1) | 0.013* |
| **MDS-UPDRS part II, mean (SD)** | 5.1<br>(6.8) | 2.3<br>(3.4) | 0.313* | 3.5<br>(4.5) | 1.7<br>(2.5) | 0.027* | 3.6<br>(4.1) | 2.4<br>(3.1) | 0.004* | 1.7<br>(3.2) | 1.6<br>(2.9) | 0.137* |
| **MDS-UPDRS part III, mean (SD)** | 8.1<br>(8.5) | 4.4<br>(4.5) | 0.052* | 4.6<br>(4.7) | 4.4<br>(5.9) | 0.696**<br>* | 5.1<br>(5.8) | 5.0<br>(5.2) | 0.077* | 3.5<br>(4.4) | 4.2<br>(5.2) | 0.727* |
| **MDS-UPDRS total score, mean (SD)** | 25.1<br>(21.0) | 14.3<br>(10.9) | 0.038* | 16.1<br>(11.8) | 14.2<br>(9.8) | 0.607** | 17.1<br>(11.2) | 14.9<br>(9.8) | 0.035** | 11.5<br>(9.5) | 11.1<br>(8.6) | 0.232* |
| **MoCA total score, mean (SD)** | 26.7<br>(2.7) | 26.4<br>(2.5) | 0.644* | 26.5<br>(2.8) | 26.6<br>(2.9) | 0.376* | 25.9<br>(2.7) | 26.5<br>(2.4) | 0.006* | 27.1<br>(2.2) | 26.9<br>(2.3) | 0.865* |
| Missing | 0 | 0 |  | 1 | 0 |  | 2 | 4 |  | 2 | 4 |  |
| **Investigator Cognitive Diagnosis, n (%)** |  |  | 0.889 |  |  | 0.987 |  |  | 0.731 |  |  | 0.447 |
| Normal cognition | 10 (83%) | 148 (83%) |  | 65 (86%) | 45 (87%) |  | 65 (92%) | 341 (87%) |  | 278 (95%) | 341 (95%) |  |
| Mild cognitive impairment (MCI) | 2 (17%) | 31 (17%) |  | 11 (14%) | 7 (13%) |  | 6 (8%) | 50 (13%) |  | 14 (5%) | 17 (5%) |  |
| Missing | 0 | 0 |  | 2 | 0 |  | 0 | 3 |  | 0 | 1 |  |
| **Cognitive Summary Score (internal), mean (SD)** | -0.33<br>(0.57) | -0.38<br>(0.71) | 0.931* | -0.26<br>(0.69) | -0.26<br>(0.75) | 0.956 | -0.23<br>(0.61) | -0.30<br>(0.73) | 0.404 | -0.06<br>(0.66) | -0.08<br>(0.65) | 0.520* |
| **SCOPA-AUT total score, mean (SD)** | 17.3<br>(10.7) | 12.8<br>(7.1) | 0.042** | 12.1<br>(8.1) | 10.9<br>(5.5) | 0.287** | 13.8<br>(8.8) | 12.2<br>(6.1) | 0.147** | 9.7<br>(6.8) | 8.7<br>(5.3) | 0.035** |
| **GDS, mean (SD)** | 3.8 (3.5) | 2.1 (2.2) | 0.081* | 3.2 (3.5) | 2.2 (2.2) | 0.578* | 3.0 (3.4) | 2.0 (2.2) | 0.159* | 1.8 (2.4) | 1.2 (1.7) | 0.003* |
| **STAI, mean (SD)** | 81.6<br>(27.3) | 63.7<br>(18.4) | 0.029* | 68.8<br>(23.4) | 64.0<br>(18.4) | 0.809* | 64.0<br>(21.4) | 60.9<br>(17.0) | 0.941* | 57.2<br>(15.6) | 54.9<br>(13.9) | 0.247* |

### Demographic and clinical characteristics vary among subtypes of early-stage NSD

We compared demographics and clinical outcomes within NSD+ cases across subgroups. Subgroups identified based on hyposmia had a higher proportion of females (Hyposmia+DEB: 36%, Hyposmia-DEB: 64%) compared with the subgroups identified based on RBD (RBD+Hyposmia: 24%, RBD-Hyposmia: 15%). UPSIT and DAT imaging were also lower in Hyposmia+DEB and Hyposmia-DEB groups compared to RBD+Hyposmia and RBD-Hyposmia groups, likely due to selection criteria (**Table 3**)

**Table 3:**
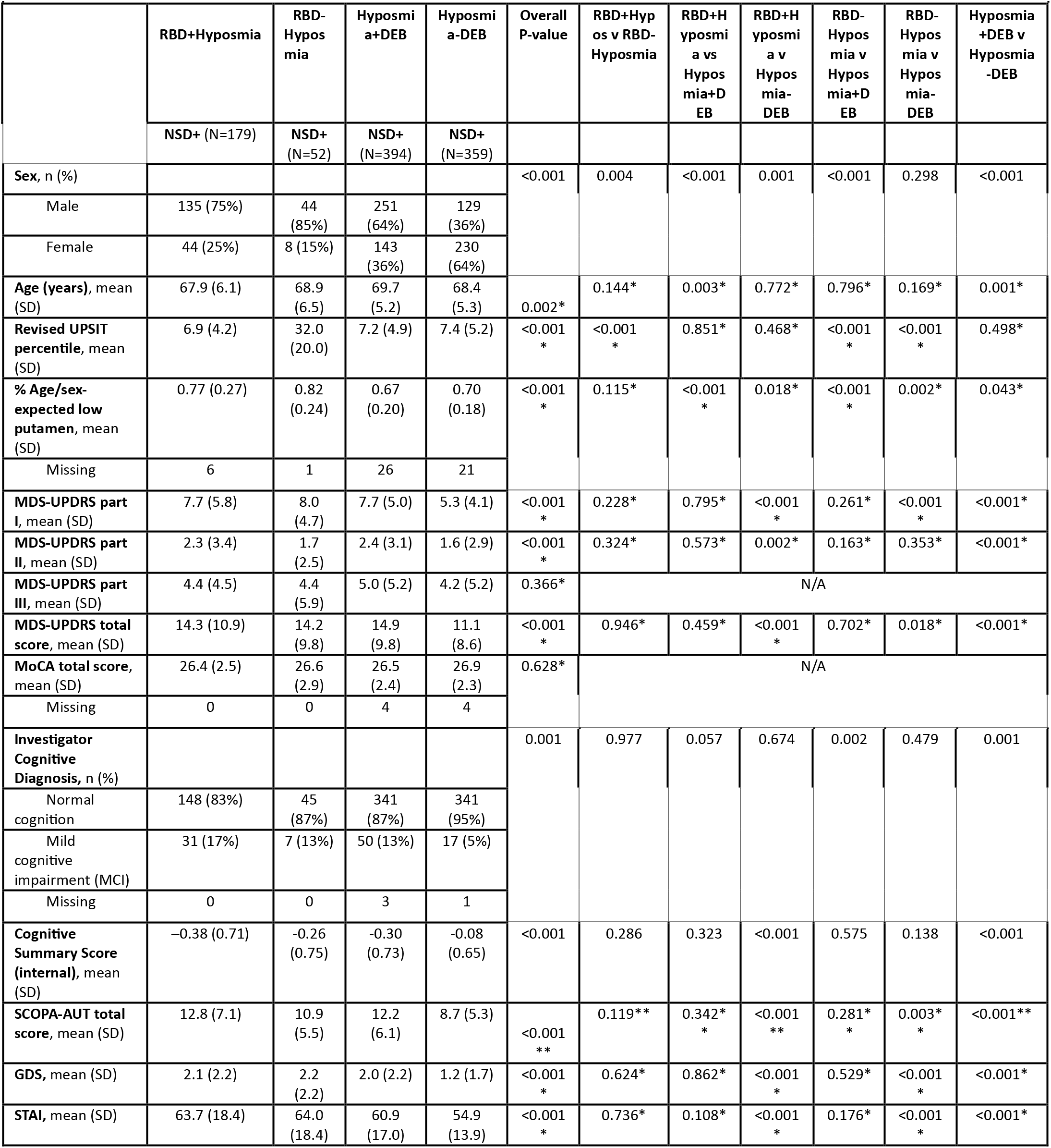
Differences in clinical assessments between people with NSD and different syndromes. All participants have positive CSFasynSAA.

As for clinical outcomes, RBD+Hyposmia and Hyposmia+DEB had higher MDS-UPDRS Parts I and II, lower cognitive summary scores and higher GDS, STAI, and SCOPA-AUT scores compared with Hyposmia-DEB participants. A higher proportion of people were determined by the investigator to have MC in the Hyposmia+DEB group (13%) compared to the Hyposmia-DEB group (5%). Participants with RBD-Hyposmia had higher MDS-UPDRS Part I, GDS, and STAI scores compared to the Hyposmia-DEB group. The MDS-UPDRS III score was similar across groups.

## Discussion

We have demonstrated that two PPMI enrollment strategies, identifying PSG-confirmed RBD from clinical study sites and central identification of hyposmic individuals, markedly enrich for positive CSFaSynSAA. The percentage of individuals positive for CSFaSynSAA varied with the recruitment strategy: RBD+Hyposmia were the most likely to be CSFaSynSAA+, followed by those with Hyposmia+DEB. Self-report of DEB with hyposmia was almost as predictive of NSD as RBD-PSG and hyposmia. Notably, individuals with RBD-Hyposmia (RBD without hyposmia) had a relatively low proportion of NSD. These findings are consistent with the range of CSFaSynSAA positivity seen in other iRBD cohorts^27^ and suggest that identifying RBD is not a sufficient enrichment paradigm for NSD, but assessment of hyposmia is also required.

In general, older people with hyposmia were also robustly enriched for CSFaSynSAA. This is consistent with prior studies showing an age-dependent enrichment of hyposmic older individuals.{Palmqvist, 2023 #203}{Marek, 2025 #559} Our finding that people with hyposmia and DEB had an increased proportion of NSD compared with Hyposmia without DEB and a similar proportion of NSD as people with RBD-PSG and hyposmia has important implications for identification of NSD+ individuals for subsequent clinical studies. DEB is readily identified with a single self-reported question.^17^ However, DEB is common – occurring in up to 19% of the general population^28^ – and only associated with PSG-confirmed RBD in a small subset,^29^ more often occurring due to other medical or psychiatric conditions. Selecting people with DEB and hyposmia enriches people who are CSFaSynSAA+ and potentially excludes people with DEB due to non-neurodegenerative reasons. Given the limited accessibility^30^ and expense^31^ of PSG, the DEB questionnaire-smell test approach provides a less expensive and more accessible way to identify these individuals who may be a relevant group with NSD to target in clinical trials aimed at prevention of disease progression. The higher overall number of participants in the Hyposmia+DEB group (n = 748) compared with the RBD-PSG group with or without hyposmia (n=386) reflects the more readily scalable nature of this approach.

Among people with NSD+, it is possible that RBD+Hyposmia and Hyposmia+DEB are a subgroup with more severe symptomatology than Hyposmia-DEB . We found that people with RBD or DEB and hyposmia have more evident autonomic, cognitive, and mood symptoms than people with hyposmia alone. We observed these differences in detailed clinical assessments as well as the investigator-determined diagnosis of MCI. Longitudinal follow-up is required to further examine the biologic and clinical progression of these individuals.

Hyposmia without DEB is likely of heterogeneous etiology, with some individuals having non-neurologic causes and some being prodromal. It may be a distinct prodromal subtype and/or may simply be earlier in the course of disease during the early NSD stages. This might explain both their lower percent of CSFaSynSAA positivity and lack of early clinical symptoms. Again, longitudinal follow-up is required to further examine the biologic and clinical progression of these individuals.

Another important finding was relatively few biologic and clinical differences between participants with NSD and non-NSD besides hyposmia. DAT deficit was consistently worse in the NSD vs non-NSD participants, but the differences were small possibly reflecting the relatively modest DAT deficit in this NSD prodromal cohort. Some clinical assessments were relatively higher (i.e, worse) in non-NSD compared with NSD. For instance, non-NSD groups had higher MDS-UPDRS Total, SCOPA-AUT, and STAI scores compared to NSD+ participants in the RBD+Hyposmia group. One explanation is that the overall scores for all participants were very low so these differences may not be meaningful. The explanation for such phenotypic differences remains to be determined with longitudinal follow up. Biomarkers and clinical characterization will be essential to address these questions.

Our study has several limitations. This was not a population-based study; individuals were enrolled based on their availability to clinical sites for RBD or their response to Smell Test direct for hyposmia. People enrolled into the RBD cohort had to have a positive PSG confirmed by the investigator, but the PSG data did not undergo centralized adjudication, which may explain the lower proportion of NSD in this group compared with other cohorts. Furthermore, Hyposmia+DEB and Hyposmia-DEB groups also had to have a DAT below 100% expected for age and sex. While this threshold is broad, it increased the CSFaSynSAA positivity in hyposmia compared with no DAT restriction. Our cohort lacks racial and ethnic diversity, and the nature of prodromal features needs to be studied in other populations.

Our biologic characterization of NSD will likely evolve to include other measures of neurodegeneration and these other pathologies may be responsible for the differences in CSFaSynSAA positivity among the prodromal groups. Longitudinal data will also help us understand whether these biologic differences among prodromal subgroups translate to faster progression of symptoms or biomarkers.

In conclusion, we demonstrate that a dual enrollment strategy of clinic-based PSG-positive RBD and centrally-identified individuals with hyposmia both develop a study population enriched for CSFaSynSAA positivity. These strategies confirm that a large biologically-derived cohort of early NSD can be identified for clinical studies. Further, we have shown that centralized recruitment offers the possibility to scale enrollment and that use of a simple screening questionnaire and smell test does identify a cohort with a high proportion of NSD. Identifying people centrally with DEB and hyposmia appears to identify a similar cohort to PSG RBD. Future studies investigating differences in underlying biology and clinical trajectories among people with early NSD with and without DEB are needed. These investigations will be important as early NSD increasingly becomes a focus for clinical trials testing therapeutics to slow progression in alpha-synuclein-related disease. Pairing a smell test with a question about DEB, a scalable screening approach, enriches cohorts not only for NSD but also for early symptomatology amenable to longitudinal follow up and intervention.

## Acknowledgments

PPMI – a public-private partnership – is funded by the Michael J. Fox Foundation for Parkinson’s Research and funding partners, including AbbVie, Alamar Biosciences, Aligning 1Science Across Parkinson’s (ASAP), Arrowhead Pharma, Arvinas, AskBio, BIAL, BioArctic, Biohaven, BlueRock Therapeutics, Bristol Myers Squibb, Calico Labs, Capsida Biotherapeutics, Critical Path Institute, DaCapo Brainscience, Denali, Edmond J. Safra Foundation, Eli Lilly, Gain Therapeutics, GE Healthcare, Genentech, GSK, Insitro, Johnson & Johnson Innovative Medicine, Lundbeck, Merck, Neumora, Neuron23, Novartis, Olink, Regeneron, Roche, Sanofi, Tenvie, UCB, Vanqua Bio, Voyager Therapeutics, The Weston Family Foundation.

## Author Contributions

EGB contributed to the conception, drafting, and design of the manuscript. HB, DEL, ARN, and NJ contributed to statistical analysis, figure and table generation, and editing of manuscript. RNA, LMC, CSC, RDD, TF, DG, KM, BM, KLP, AS, CMT, DW contributed to the editing of the manuscript. KM, TS, TFT contributed to conception, editing, and design of the manuscript.

## Data Availability

Data used in the preparation of this article were obtained on 2025-06-09 from the Parkinson’s Precision Medicine Initiative (PPMI) database (https://www.ppmi-info.org/access-data-specimens/download-data), RRID:SCR_006431. For up-to-date information on the study, visit http://www.ppmi-info.org.

This analysis was conducted by the PPMI Statistics Core and used actual dates of activity for participants, a restricted data element not available to public users of PPMI data.

Protocol information for The Parkinson’s Precision Medicine Initiative (PPMI) Clinical - Establishing a Deeply Phenotyped PD Cohort can be found on protocols.io or by following this link: https://dx.doi.org/10.17504/protocols.io.n92ldmw6ol5b/v2.

## Code Availability

Statistical analysis codes used to perform the analyses in this article are shared on Zenodo [https://doi.org/10.5281/zenodo.21534825].

